# Dose-Response Relationships between Physical Exercises and Mental Health during Early Adolescence: an Investigation of the Underlying Neural and Genetic Mechanisms from the ABCD Study

**DOI:** 10.1101/2023.09.20.23295830

**Authors:** Gechang Yu, Xinran Wu, Zhaowen Liu, Mai Shi, Huaxin Fan, Yu Liu, Nanyu Kuang, Songjun Peng, Zhengxu Lian, Jingyang Chen, Senyou Yang, Chuiguo Huang, Hongjiang Wu, Baoqi Fan, Jianfeng Feng, Wei Cheng, Barbara J. Sahakian, Trevor W. Robbins, Benjamin Becker, Jie Zhang

## Abstract

Adolescence is a critical developmental with increased vulnerability to mental disorders. While the positive impact of physical exercise on adult mental health is well-established, dose-response relationships and the underlying neural and genetic mechanisms in adolescents remain elusive. Leveraging data from >11,000 pre-adolescents (9-10 years, ABCD Study) we examined associations between seven different measures of exercise dosage across 15 exercises and psychopathology, and the roles of brain function and structure and psychiatric genetic risks. Five specific exercises (basketball, baseball/softball, soccer, football, and skiing) were associated with better mental health while the beneficial effects varied with exercise types, dosage measures and dimensions of psychopathology. Interestingly, more exercise does not always translate to better mental health whilst earlier initiation was consistently advantageous. Communication between attention and default-mode brain networks mediated the beneficial effect of playing football. Crucially, exercise mitigates the detrimental effects of psychiatric genetic risks on mental health. We offer a nuanced understanding of exercise effects on adolescent mental health to promote personalized exercise-based interventions in youth.

## Introduction

Adolescence is a key period for significant physical and neurocognitive development. Individual physical exercise (PE) and physical activity (PA) habits and preference for specific exercise types tend to develop during this period and remain stable during later life (*1–3*). In parallel, most mental disorders also originate in adolescence with consequences that reach far into adulthood (*4, 5*). Increasing evidence supports the benefits of PE on mental health, including reducing anxiety (*6, 7*) and depression (*8, 9*) as well as promoting general resilience across the lifespan (*10*). Therefore, exercise-based interventions have been proposed as an alternative to conventional pharmacotherapies in mental disorders (*11*). Similar to other interventions such as pharmacological agents (*12, 13*), exercise dosage (e.g., frequency, duration and amount) seems to crucially contribute to both, promotion of beneficial and avoidance of adverse effects. However, while dose-response relationships have been extensively examined for pharmacological interventions, little is known currently about how exercise dosage, such as exercise type, duration, frequency and intensity, are related to mental health, especially during childhood and adolescence and it remain a matter of debate whether more exercise is always better (*14*). In addition, little is known about the optimal age for children and adolescents to start participating in PEs in order to obtain psychological resilience.

Previous relevant studies (*14–16*) primarily focused on large categories of exercises, such as aerobic vs resistance exercises, or team vs individual sports, rather than specific kinds of organized exercises. Considering that adolescents often have specific preferences for different kinds of exercises, it is also important to investigate whether different kinds of PEs have distinct effects and mechanisms. For example, basketball, baseball and soccer all belong to team sports but require quite different motor and cognitive skills, and in turn may not have the same effects on mental health. In brief, determining the dose-response relationships of different PEs is a crucial first step that can help clinicians to prescribe precise and personalized exercise interventions based on the youth’s interest as well as mental health profile.

Although the neurobiological mechanisms underlying the effects of PE on mental health still remains unclear, several hypotheses have been proposed. For instance, participation in exercise could increase the level of brain-derived neurotrophic factor, reduce the level of inflammation, improve synaptic plasticity (*17, 18*) and increase hippocampal volume (*18–20*) and can thus promote age- and duration-dependent changes in brain activity (*21*). However, it remains unclear whether different types and dosage measures of exercises affect mental health through different neural pathways (*22, 23*). Besides, since mental health is influenced by both genetic and environmental (e.g., lifestyle) factors, it is important to identify how interactions between PEs and genetic risks of psychopathology influence mental health, which could render individualized PE recommendations as an efficacious early intervention to mitigate an increased psychiatric genetic risks.

In the current study (Figure 1), we utilized a large-scale (n=11,878) multisite population-representative adolescent (aged 9-10 years at baseline) cohort from the US, the Adolescent Brain and Cognitive Development (ABCD) Study^®^ (*24*), which includes 23 types of PEs and corresponding dosage measures, mental health measures, multimodal neuroimaging measures and genotype data. We aim to investigate (1) the dose-response relationships between different types of PEs and adolescent mental health; (2) if the beneficial effects of dosage of PEs on mental health are mediated by variations in the functional and structural architecture of the brain, and (3) how the interactions between PE and psychiatric genetic risks influence mental health.

**Figure 1.**
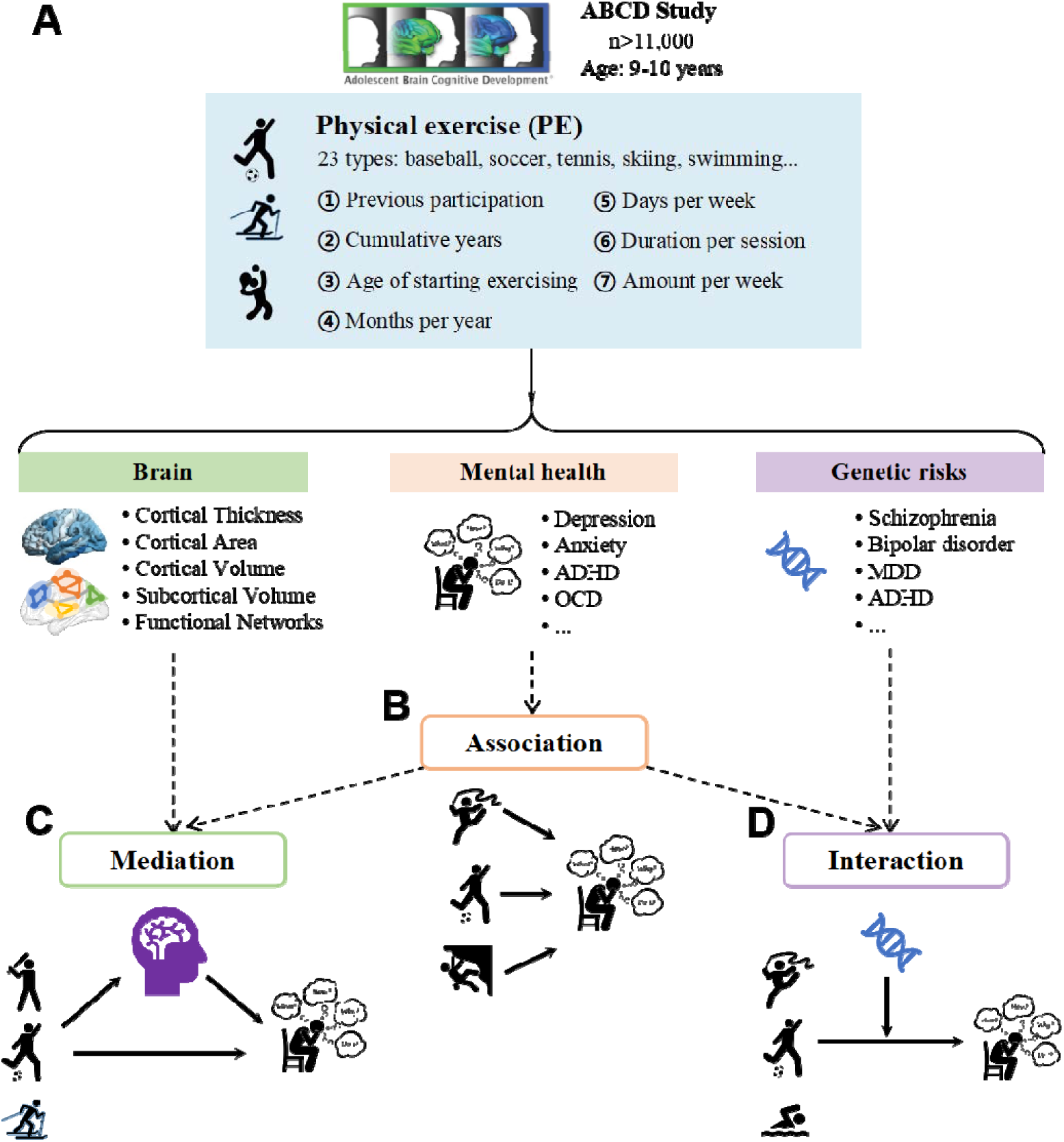
Overview of the study design and analyses. **(A)** The multimodal data used in our study was from the ABCD study, which includes over 11,000 US adolescents (9-10 years). The data of physical exercises (PEs) includes 23 types and each type of PE has seven measures of dosage. The brain imaging data includes structural and functional measures. Mental health data includes assessment of multiple dimensions of mental and behavioral problems for youths from CBCL scales (Child Behavior Checklist). Youths were also assessed for their genetic risks of psychopathology using large-scale GWAS (Genome-Wide Association Study) summary data. **(B)** For each type of PE, we examined the associations between seven dosage measures of PE and mental health. **(C)** For each type of PE with ‘beneficial’ effects, we examined if neural correlates related to PE dosage mediated the beneficial effects of PE on mental health. **(D)** We assessed how the interactions between psychiatric genetic risks and PE affect mental health.

## Results

### Characteristics of physical exercises in ABCD

Participation and dosage of 23 types of PEs were assessed using Parent Sports and Activities Involvement Questionnaire (*25*) in ABCD release 4.0. We excluded PEs with small number of participating youths (n<400) and focused on the 15 most common kinds of PEs (refer to Table 2 and Table S1 for demographics). From the parent-reported questionnaire, we utilized seven measures ranging from a larger time scale (previous participation, cumulative years and age of starting exercising) to a smaller time scale (months per year, days per week, duration per session, amount per week) to quantify the dosage of PE (Figure 1).

Soccer (41%) represented the most popular PE among US youths from the ABCD study. The proportions of participation in any sports is very similar in boys (85.7%) and girls (83.1%) while the proportion varies between different kinds of sports (Table 2 and Supplemental Table S1-2). The proportion of Black youths participating in any sports (69.7%) is comparatively lower than in White (90.8%) and Asian (88.9%) youths. As to the socio-economic status (Supplemental Table S1), the higher household income and higher parental education, the higher the proportion of youths’ participation in any sports. In different BMI groups (Supplemental Table 1), severely underweight youths had the lowest proportion (71.1%) of participating in any sports while the proportions of participating in any sports in overweight (83.8%) and obese (77.9%) youths were close to that in normal weight youths (87.3%). Besides, most youths (94.7%) have participated in sports organized in teams or groups (Supplemental Table S2). Importantly, participation in different kinds of sports generally did not show significant associations between each other (for visual presentation see Figure S1). There were only weak correlations among team ball sports and between ballet/dance and gymnastics.

### Dose-response relationships between PE and mental health

We first compared dimensional psychopathology, including Total, Externalizing, Internalizing and Thought Problems scores from the Child Behavior Checklist (CBCL), between youths with and without previous participation in each type of PE as well as any types of PE using linear mixed model (LMM) and adjusted for age (except for age of starting exercising), sex, race, household income, parental education, pubertal level, body max index (BMI), and random effects for family nested within site. A two-side p<0.001 was set as the significance threshold. As Figure 2A shows, youth’s participation in any sports was significantly (p<0.001) associated with lower CBCL Total Problems (beta=-2.00, se= 0.48, z=-4.17, p=3.1×10^-5^), Internalizing Problems (beta=-0.58, se=0.15, z=-3.85, p=1.2×10^-4^) and Thought Problems (beta=-0.21, se=0.06, z=-3.58, p=3.3×10^-4^). However, specific types of PEs exhibited distinct association patterns (Figure 2A): participation in baseball/softball, basketball, soccer, football and skiing were significantly (p<0.001) associated with reduced Total Problems or other dimensions of psychopathology while participation in martial arts was significantly (p<0.001) associated with increased Total Problems as well as Externalizing, Internalizing and Thought Problems. Other types of sports did not exhibit significant (p<0.001) associations with mental health burden. Results were similar for other CBCL scales (Figure S2). Therefore, for youths participating in each type of ‘beneficial’ sports (baseball/softball, basketball, soccer, football and skiing), we ask the question if more exercise and earlier start in participation is better for mental health.

**Figure 2.**
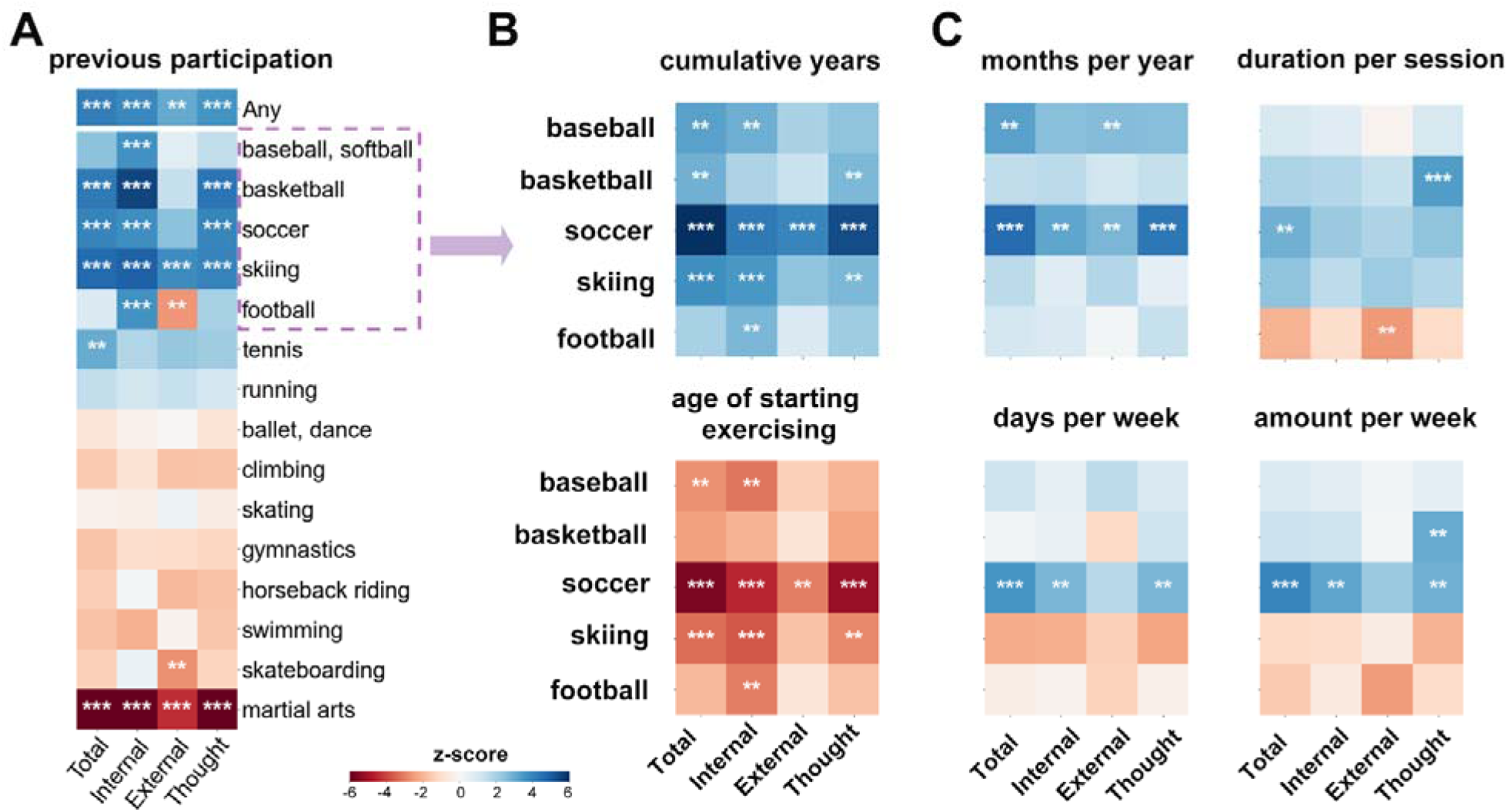
The relationships between seven dosage measures of physical exercises and CBCL scores. **(A)** The relationships between previous participation of any PE and one of the 15 PEs and CBCL Externalizing, Internalizing, Thought and Total Problems scores. **(B)** The relationships between cumulative years and age of starting exercising of five ‘beneficial’ PEs and CBCL Externalizing, Internalizing, Thought and Total Problems. **(C)** The relationships between months per year, days per week, duration per session and amount per week of five ‘beneficial’ PEs and CBCL Externalizing, Internalizing, Thought and Total Problems. The colorbars in **(A-C)** represent the z-score of the regression coefficient from LMM. The asterisks in **(A-C)** indicate: **, p<0.01; ***, p<0.001.

Firstly, we examined the associations between six detailed dosage measures (cumulative years, age of starting exercising, months per year, days per week, duration per session and amount per week) in these five kinds of ‘beneficial’ PEs and four dimensions of psychopathology. We found that these dose-response relationships for ‘beneficial’ PEs varied in different dosage measures (Figure 2B-C). On the one hand, for cumulative years and age of starting exercising (Figure 2B), it is notable that longer cumulative years and earlier start in most ‘beneficial’ PEs were significantly (p<0.001) associated with better mental health, especially for soccer and skiing. For example, earlier participation in soccer was significantly associated with lower Total Problems (beta=0.77, se=0.14, z=5.64, p=1.8×10^-8^) as well as Internalizing (beta=0.20, se=0.04, z=4.56, p=5.4×10^-6^) and Thought Problems (beta=0.09, se=0.02, z=5.17, p=2.4×10^-7^). Baseball/softball and basketball also exhibited similar directions though did not pass the significance threshold. On the other hand, for the dosage measures on a more fine-grained time scale (Figure 2C), the association patterns were quite similar. For months per year, days per week and amount per week, only soccer showed significant (p<0.001) associations with better mental health. Months per year (beta=-0.41, se=0.09, z=-4.64, p=3.7×10^-6^), days per week (beta=-1.00, se=0.28, z=-3.53, p=4.2×10^-4^) and amount per week (beta=-0.62, se=0.16, z=-3.96, p=7.6×10^-5^) of soccer were all associated with lower Total Problems. For duration per session, only basketball was significantly associated with lower Thought Problems (beta=-0.09, se=0.03, z=-3.36, p=7.9×10^-4^).

Secondly, to further validate the linear relationships between PE dosage measures and mental health burden identified for the 5 beneficial exercises, and examine if there were nonlinear relationships (J-shaped or U-shaped) between them, we (1) categorized each dosage into several groups ranging from low to high levels, (2) compared mental health burden differences between low level group and other groups and (3) tested if there existed linear trend relationships across groups. Specifically, months per year is categorized as 1-3 months, 4-6 months, 7-9 months and 10-12 months. Days per week is categorized as ≤1 day, 2-3 days, 4-5 days and >5 days. The duration per session of each PE is divided into <30 min, 30-60 min, 60-90 min, 90-120 min and ≥120 min. The amount per week of each PE is divided into <1 hour, 1-2 hours, 2-3 hours, 3-4 hours and >4 hours. Age of starting exercising is divided into toddler (<3 years), preschooler (3-5 years), middle childhood (6-8 years) and early adolescence (9-10 years).

The results were generally consistent with the linear relationships identified in the first step and we did not observe any significant nonlinear relationships like (inverted) J-shaped or (inverted) U-shaped ones. On the one hand, for age of starting exercising (Figure 3), there were also significant linear trend relationships between most beneficial PEs and CBCL Total Problems, which further suggested earlier start participation exercise might be better for mental health. For months per year, days per week and amount per week (Figure S3), only soccer exhibited significant linear trend relationships with CBCL Total Problems. For duration per session, we also found significant (p(trend)=4.6×10^-4^) linear trend relationships between basketball and CBCL Thought Problems (Figure S4). Therefore, except for playing soccer and basketball, more exercise might not produce additional benefits for mental health. In other words, lower dose level might be sufficient to obtain beneficial effects.

**Figure 3.**
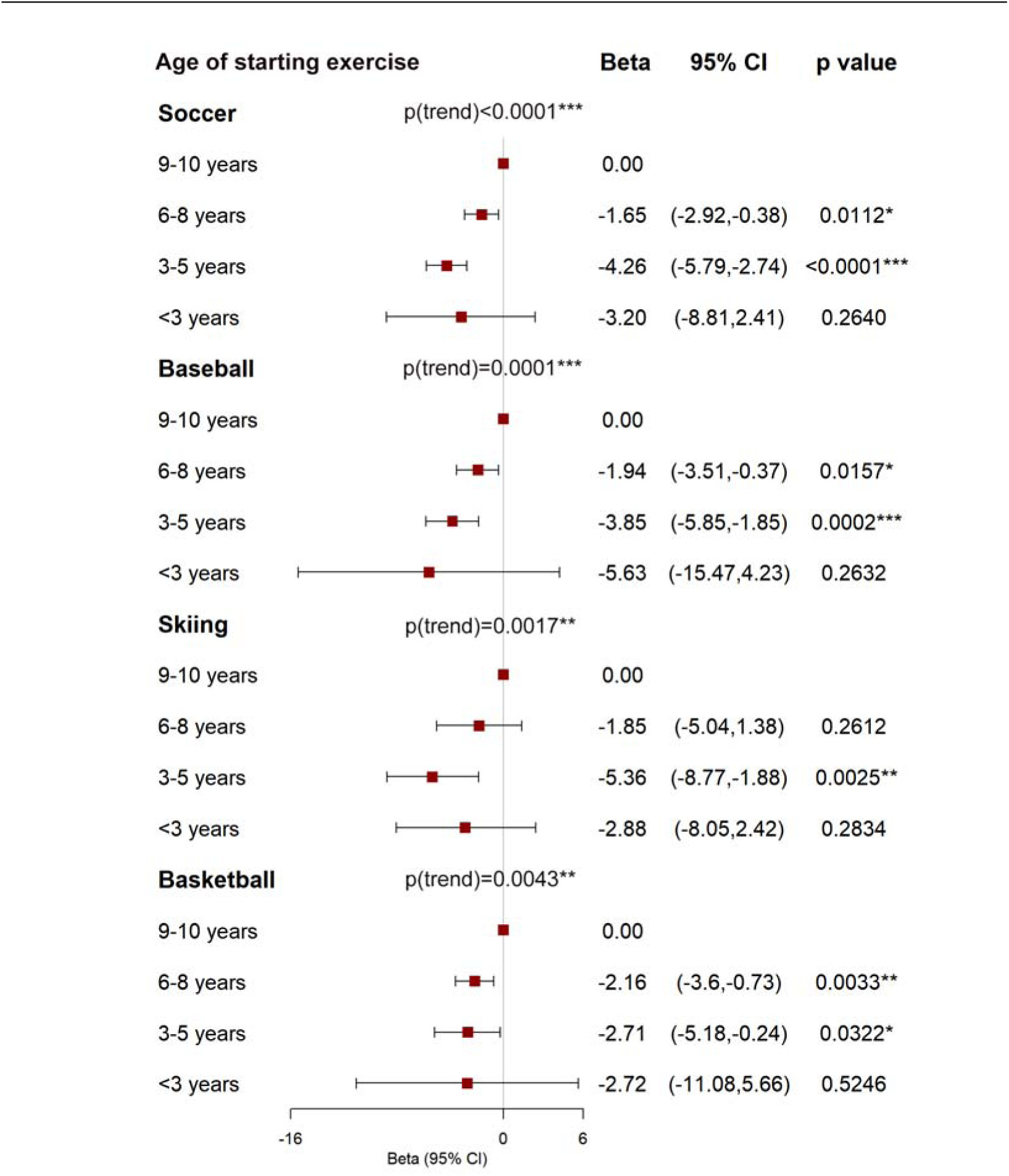
The associations between different periods of age of starting participation in soccer, baseball, skiing and basketball and CBCL Total Problems. Betas were adjusted for sex, race, household income, parental education, pubertal level, body max index (BMI), and random effects for family nested within site. p(trend) means p value for linear trend test. *: p<0.05; **: p<0.01; ***: p<0.001.

### Neural correlates of exercise dosage

To investigate the structural and functional brain architecture underlying these dosage measures of ‘beneficial’ exercises, we examined the associations between seven dosage measures and brain morphology, including cortical thickness (CT), area (CA), volume (CV) and subcortical volume (subCV), and functional network measures, including resting-state functional connectivity between and within cortical networks, and between cortical and subcortical structures. We found that different types of ‘beneficial’ PEs exhibited both common and specific neural correlates. For previous participation (Figure 4B), playing baseball was associated with increased functional connectivity (beta=0.01, se=0.003, z=3.43, p=6×10^-4^) between retrosplenial temporal network (RSP) and salience network (SAN). Playing football was associated with increased functional connectivity between dorsal attention network (DAN) and ventral attention network (VAN) (beta=0.007, se=0.002, z= 3.34, p=8.3×10^-4^), between DAN and default mode network (DMN) (beta=0.007, se=0.002, z= 3.41, p=6.4×10^-4^) and within sensorimotor hand network (SMH) (beta=0.01, se=0.003, z= 3.96, p=7.6×10^-5^) as well as with decreased connectivity between DAN and visual network (VIS) (beta=-0.009, se=0.002, z=-3.73, p=1.9×10^-4^) and between fronto-parietal network (FPN) and SMH (beta=-0.007, se=0.002, z=-3.41, p=6.6×10^-4^). Skiing was associated with decreased connectivity between DAN and auditory network (AUD) (beta=-0.1, se=0.002, z=-3.97, p=7.5×10^-5^).

**Figure 4.**
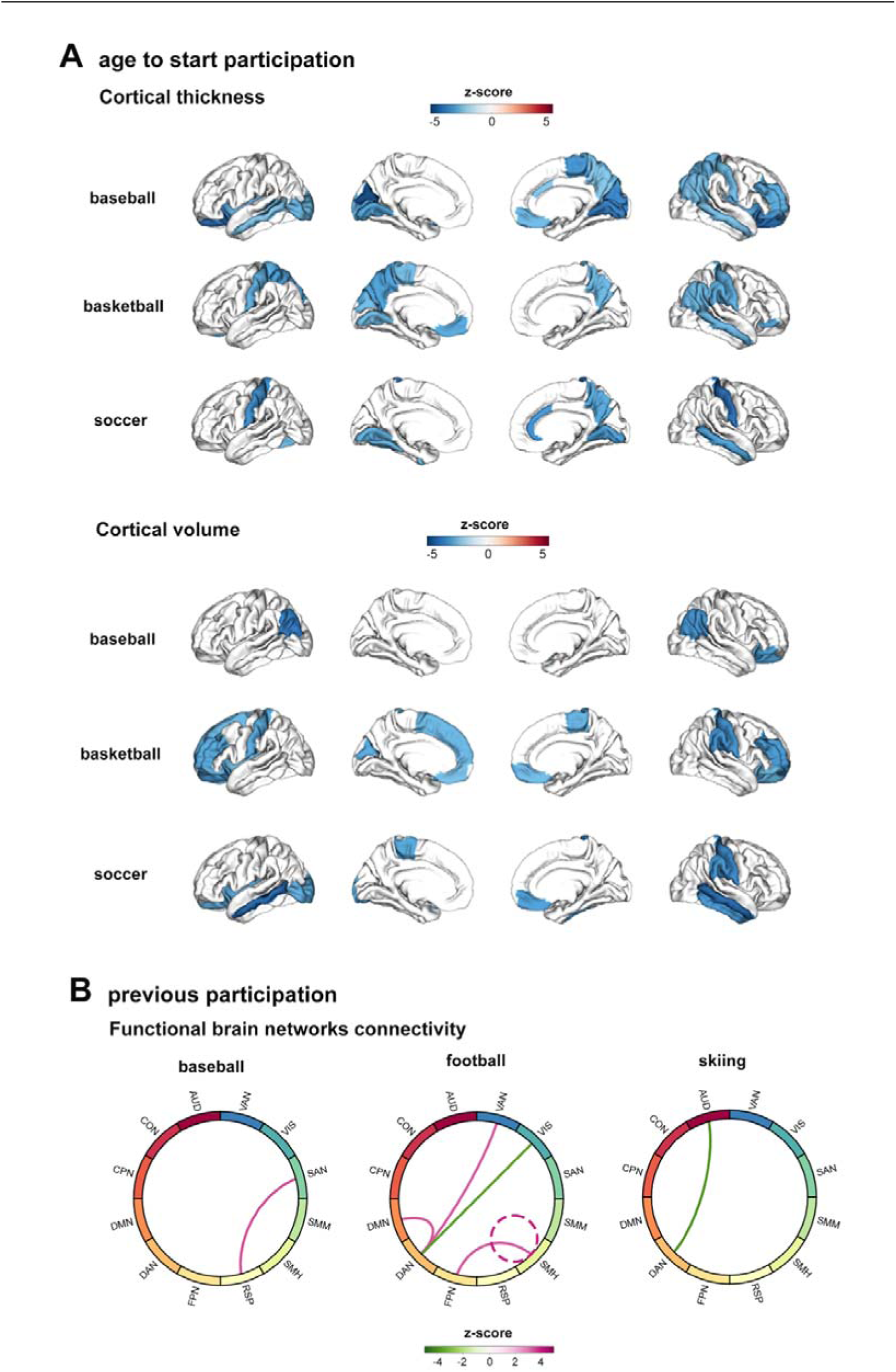
Neural correlates of dosage measures of those ‘beneficial’ PEs. **(A)** Cortical thickness and cortical volume significantly (P_FDR_<0.05) associated with age of starting participation in baseball, basketball and soccer. **(B)** Functional connectivity between and within functional networks significantly (P_FDR<0.05_) associated with previous participation of baseball, football and skiing. The colorbars in **(A)**-**(B)** represent the z-score of the regression coefficient from LMM. Abbreviations: AUD=auditory network, CON=cingulo-opercular network, CPN=cingulo-parietal network, DMN=default mode network, DAN=dorsal attention network, FPN=fronto-parietal network, RSP=retrosplenial temporal network, SMH=sensorimotor hand network, SMM=sensorimotor mouth network, SAN=salience network, VIS=visual network, VAN=ventral attention network.

For age of starting exercising, an earlier start in playing baseball, basketball and soccer were consistently associated with significantly increased CT (Figure 4A), with shared increased CT in visual, visual association and somatosensory cortex, including left lingual gyrus, right middle temporal gyrus, right postcentral gyrus and right precuneus. Earlier start of playing baseball, basketball and soccer was also significantly associated with significantly increased CV in lateral orbitofrontal cortex (Figure 4A). For days per week, playing basketball was also associated with increased CT across multiple regions (Figure S5). For duration per session (Supplemental Table S3), we found duration of playing basketball was associated with increased functional connectivity between RSP and DMN (beta=0.004, se=0.001, z=3.39, p=7.2×10^-4^) and between RSP and FPN (beta=0.003, se=0.001, z=3.27, p=0.001). Duration of skiing was significantly associated with reduced functional connectivity between DAN and right thalamus (beta=-0.007, se=0.002, z=-3.76, p=1.9×10^-4^) and between FPN and brain stem (beta=-0.007, se=0.002, z=-3.77, p=1.8×10^-4^). Please refer to Supplemental Table S3 for other significant neural correlates.

### Mediator role of neural correlates on the dose-response relationships

To further explore whether PE-related neural correlates mediated the beneficial effects of PE on mental health, we performed Mediation analyses with 10,000 bootstraps. We found that the functional connectivity between DAN and DMN significantly mediated the beneficial effects of football on Internalizing Problems (β [95% CI] =0.026 [0.009, 0.052], p= 8.7×10^-5^), Depressive Problems (β [95% CI]=0.010 [0.004, 0.020], p= 9.6×10^-5^) and Withdrawn/Depressed (β [95% CI]=0.010 [0.004, 0.019], p= 1.0×10^-4^) (Figure 5A and Figure S6). In addition, functional connectivity between DAN and VAN significantly mediated the beneficial effects of participation in football on Depressive Problems (β [95% CI]=0.009 [0.003, 0.019], p= 9.1×10^-5^) and Internalizing Problems (β [95% CI]=0.022 [0.007, 0.045], p= 4.4×10^-4^) (Figure 5B and Figure S6).

**Figure 5.**
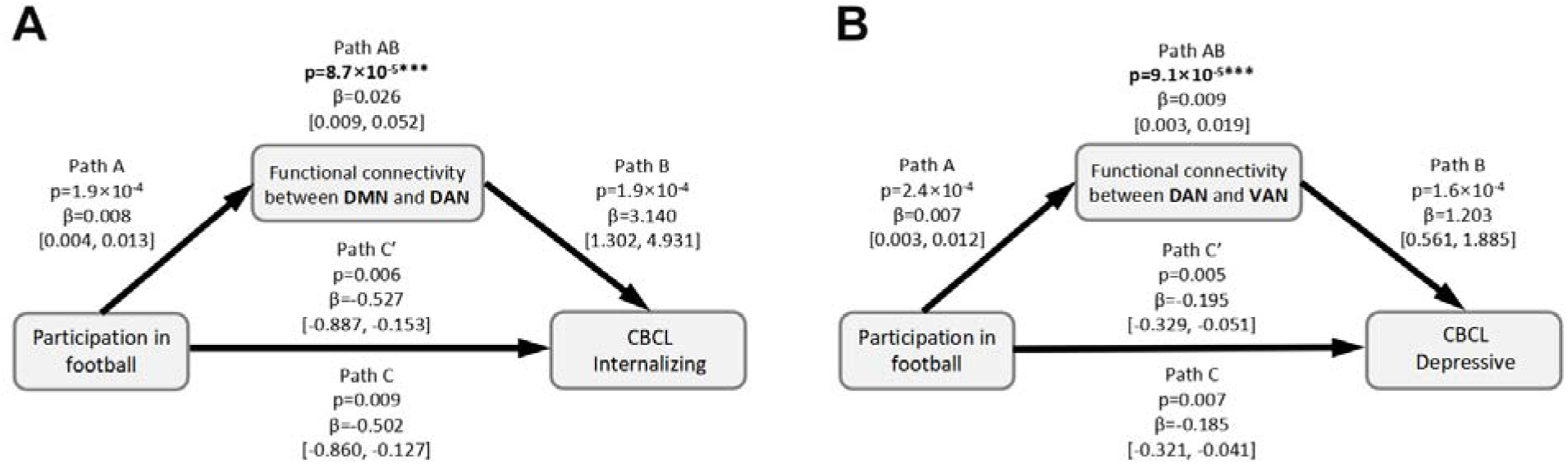
Mediation analysis. **(A)** Functional connectivity between default mode network (DMN) and dorsal attention network (DAN) mediates the beneficial effect of participation in football on CBCL Internalizing Problems. **(B)** Functional connectivity between dorsal attention network (DAN) and ventral attention network (VAN) mediates the effect of participation in football on CBCL Depressive Problems. The 95% confidence intervals are listed in the bracket. In **(A-B)**, Path AB represents the average causal mediation effects (ACME). Path C represents the total effect without mediator. Path C’ represents the direct effect accounting for the indirect effect of mediator. The asterisks in **(A-B)** indicate: *, p<0.05; **, p<0.01; ***, p<0.001.

### Genetic risk-buffering role of exercise on mental health

To explore how the interactions between genetic risks of psychopathology and PEs may influence mental health, we also assessed the effects of interactions between ten polygenic risk scores (PRSs) for various psychiatric disorders and exercise dosage on mental health. Psychiatric disorders and associated traits here include schizophrenia (SCZ) (*26*), psychotic experiences (PSE) (*27*), post-traumatic stress disorder (PTSD) (*28*), bipolar disorder (BIP) (*29*), major depressive disorder (MDD) (*30*), depression (*31*), neuroticism (*31*), obsessive-compulsive disorder (OCD) (*32*), Attention-Deficit/Hyperactivity Disorder (ADHD) (*33*) and risk tolerance (*34*),

We observed that participation in any physical exercises could significantly buffer the genetic risks of ADHD on Total Problems, Social Problems, Attention Problems and thought Problems (P_FDR_<0.05, Figure 6A). In other words, among youths with increased genetic risk of ADHD, youths with participation in any sports exhibited a significantly lower mental health burden, especially Thought Problems (beta=-0.90, se=0.27, z=-3.39, p=7.2×10^-4^), than those who do not exercise at all (Figure 6C). Besides, duration per session and amount per week of playing baseball also buffered the effects of PSE PRS on mental health burden (Figure S7). However, we also observed that months per year of playing football could amplify the effects of PRS for risk tolerance on Externalizing Problems, Conduct Problems and Rule-breaking behavior (Figure 6B) which may reflect a highly complex interaction between genetic risk, dosage, sports and other environmental factors.

**Figure 6.**
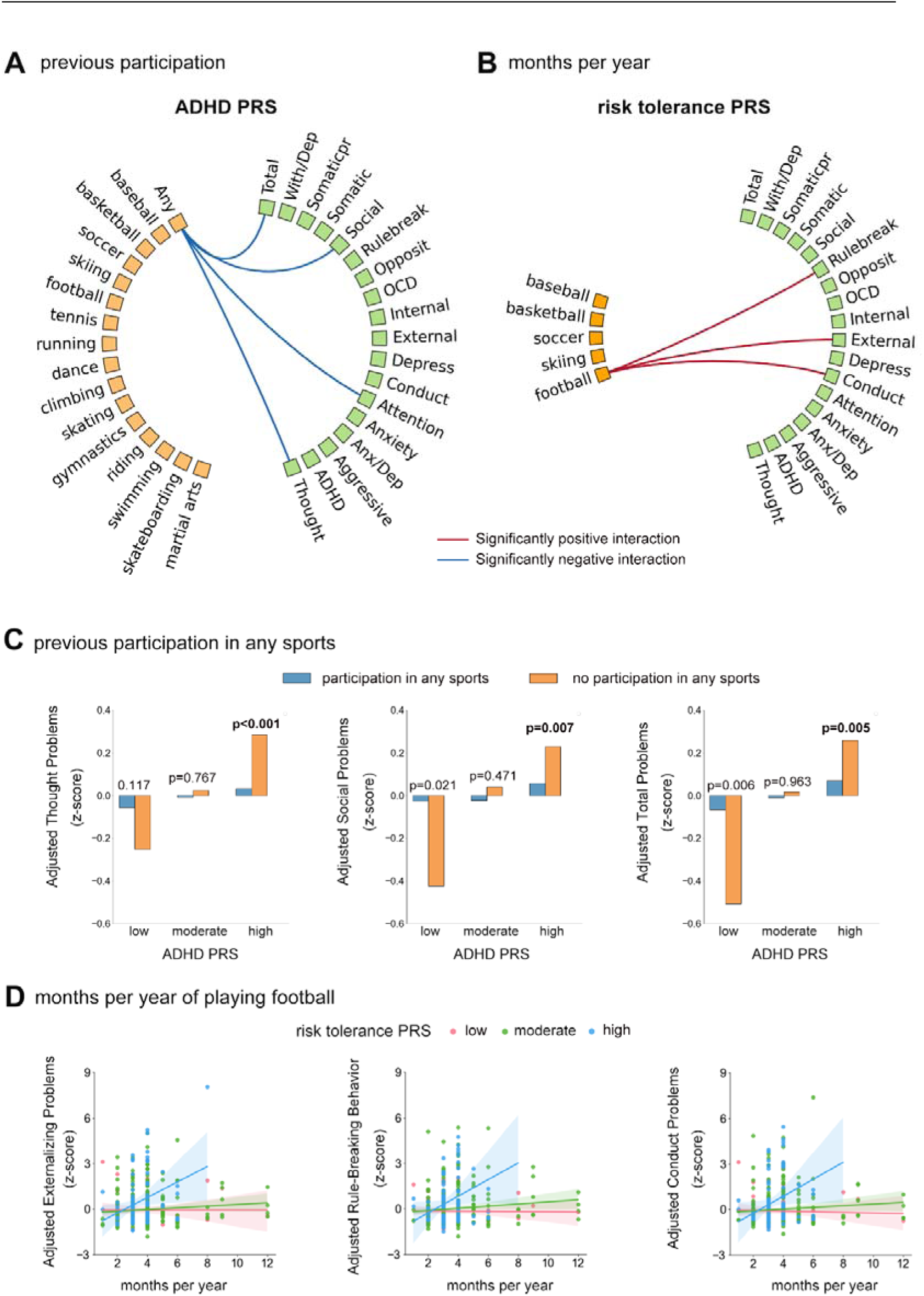
The effects of the interactions between psychiatric genetic risks and physical exercises on mental health. **(A)** The effects of ADHD PRS on the associations between previous participation and mental health. **(B)** The effects of risk tolerance PRS on the associations between months per year of five ‘beneficial’ exercises and mental health. **(C)** Differences of mental health burden between youths participating in any sports and youths without any participation in youths with low, moderate and high ADHD PRS. **(D)** The associations between months per year of playing football and mental health burden in youths with low, moderate and high risk tolerance PRS. X axis: groups of low, moderate and high ADHD PRS. Y axis: CBCL scales adjusted for covariates. The red line in **(A-B)** indicates significantly (P_FDR_<0.05) positive interaction between ADHD PRS and physical exercises while the blue line in **(A-B)** indicates significantly (P_FDR_<0.05) negative interaction between ADHD PRS and physical exercises. The p value in **(C)** indicates the difference in mental health burden between youths with or without participation in any sports.

## Discussion

Our study provided a comprehensive and fine-grained profile of the dose-response relationships between 7 dosage measures (previous participation, cumulative years, age of starting exercising, months per year, days per week, duration per session and amount per week) of PEs and dimensions of adolescent mental health in a population-representative cohort including over 11,000 adolescents. Participation in five types of common PEs were significantly associated with better mental health, including baseball/softball, basketball, soccer, football and skiing. However, this beneficial effect varied as a function of dosage measure, type of exercise and dimension of psychopathology. Interestingly, more exercise did not always exert more beneficial effects on mental health outcomes while starting exercise earlier consistently showed a beneficial effect on mental health and may promote the development of resilience. Our study further highlighted the important role of brain architecture in general and of PE specific pathways in particular on the positive effects of PE on mental health in youths, such that the functional connectivity between DAN and DMN as well as VAN crucially mediated the beneficial effects of playing football on internalizing problems, which shed light on the neural pathways between PE and mental health. Importantly, our genetic risk analyzes demonstrated that engagement in PE could buffer the detrimental effects of elevated psychiatric genetic risks on mental health during early adolescence.

While participation in in any sports showed significant associations with better mental health, we observed a considerable heterogeneity in the effects of different types of PE on mental health, such that not all sports were associated with better mental health. We found both specific types of team sports (baseball/softball, basketball, soccer, football and skiing) and individual sports (skiing and tennis) showed associations with better mental health. Previous studies (*14, 15, 22, 35*) have shown positive associations between team sports and mental health. However, Kunitoki et al. (*15*) and Hoffmann et al. (*35*) also found that individual sports were significantly associated with worse mental health in the ABCD Study. This inconsistency is possibly due to the heterogeneity of effects among different types of individual sports. Different sports have both similar and different motor patterns and specific needs for power, coordination, endurance, balance, agility and speed (*36*), which may in turn promote distinct effects on mental health. Therefore, it is also important to parse out the effects of specific types of PEs to pinpoint the similarity and difference between them, which would otherwise be obscured in the larger categories (team and individual sports). Besides, these PE-specific ‘beneficial’ effects also operated on specific dimensions of psychopathology. For example, only skiing was significantly associated with lower Externalizing Problems while baseball/softball and football were only significantly associated with lower Internalizing Problems. Therefore, different types of sports may have different effects on different dimensions of psychopathology. Prior studies focus on single aspect of mental illness like depression rather than taking full dimensions of psychopathology into account (*37*). In brief, our findings may allow clinicians to make more personalized recommendation of sport interventions to youth, which are tailored to their interest and different dimensions of mental health.

Accumulating evidence from recent studies suggests that more exercise is not always better for mental health in adults (*14, 38*). Our results filled a current research gap in adolescents and further validated this emerging perspective. For frequency, duration and amount of PEs, not all ‘beneficial’ sports showed significant dose-response (linear or nonlinear) relationships with mental health. In other words, for some types of ‘beneficial’ exercises like baseball/softball, skiing and football, higher frequency, duration and amount may not necessarily produce more beneficial effects while lower level of dose are sufficient to obtain beneficial effects. Our results were consistent with Chekroud et al. (*14*) and Harvey et al. (*38*), who also found that excessive engagement in exercise did not produce any additional benefit on mental health in adults and may even produce adverse effects. Our results highlighted that an earlier initiation of participation in exercise could be a protective factor for youths’ mental health. Previous studies have linked early participation in sports with better physical fitness and psychological well-being and self-esteem (*39, 40*). We found that preschool age (3-5 years) or middle childhood (6-8 years) might be the optimal age to start playing soccer, baseball/softball, basketball and skiing for better mental health. Preschool age and middle childhood are important development periods during which physical, social and mental skills rapidly develop and children start to move into expanding roles and environments. Starting to participate in physical exercise during this period, especially team sports, could help children achieve new motor and social skills promote confidence thus facilitate cognitive development.

Our study systematically investigated the relationships between dosage of exercise and brain structural and functional architecture during early adolescence. We found both common and specific neural pathways associated with dosage of different exercises. For instance, earlier starting participation in baseball, basketball and soccer shared increased CT in visual, visual association and somatosensory cortex, which may undergo an earlier cortical thinning compared to other regions (*41*). This result suggests that early participation in exercise could attenuate the extent of cortical thinning in these areas. While previous studies have linked PA with increased hippocampal volume (*16*) and white matter microstructure integrity (*42*) in youth, little is known about how PE/PA affects the functional communication and interaction among cortical brain networks and between cortical and subcortical structures. Our results suggested that different types of sports exhibit distinct patterns of influence on brain functional architecture. Taken together, these distinct neural correlates (structural and functional) for each type of PE are key to understand the neurobiological underpinnings of differences (e.g., motor and social skills) between different types of sports. More importantly, our study highlighted the mediating role of functional connectivity between DAN and VAN and between DAN and DMN in the beneficial effects of playing football on internalizing problems. The DAN closely interacts with the VAN and DMN, which was involved in top-down and bottom-up attentional control during daily life (*43, 44*). Abnormal functional communication between these networks has been implicated in MDD (*45*) and anxiety (*46*) and attentional bias and selective attention to potential threats represent candidate pathomechanisms involved in depression and anxiety (*47*). Therefore, playing football may promote youths’ attentional control through strengthened functional integration between DAN, VAN and DMN, which in turn may normalize the attentional bias and improve the internalizing problems.

Physical and mental health are influenced by both genetic (e.g., genes and epigenetics) and environmental factors (e.g., living conditions, pollution, lifestyle and socioeconomic factors). Early life represents a critical period to to illuminate how the interactions between environmental and genetic factors affect health, which might be less detectable and modifiable in later life. The Gene–Environment (G × E) interaction analysis in our study showed that physical exercises could buffer or offset a considerable amount of the detrimental effects of a genetic burden of psychopathology, especially for ADHD and PSE, on mental health during early adolescence. Kunitoki et al. (*15*) also found participation in team sports could buffer the effects of ADHD PRS on mental health in ABCD study though they focused on large categories of team sports, individual sports and nonsports activities. The risk-buffering role of PE has important implications for public health and clinical interventions: youths with a high genetic risk for mental disorders could be encouraged to participate in PE to strengthen resilience to the adverse effects of genetic risks on mental health during this developmentally vulnerable period. Further, our study also underlined the potential advantage of genetic risk profiling, which might help early screen youths with elevated genetic risk of psychopathology and tailor exercise programs for them.

Finally, our study showed inequalities of physical exercise participation in US youths; youths from families of lower socioeconomic status and of black race engaged in less physical exercise, which has also been documented before (*48*). Lack of facilities and resources, high costs, less support and instruction from families and communities could be barriers for them to have equal opportunities to participate in physical exercise, especially organized sports (*48*). Therefore, it is necessary for the government and communities to provide more support and resources for black youths and youths living with lower socio-economic status to increase their opportunities to participate in physical exercise, which will contribute to their physical and mental health.

We acknowledge that there are several limitations in our study. First, we did not further explore why some sports (e.g., martial arts) were associated with higher level of psychopathology. Other unmeasured factors and bidirectional causal relationships (*49*) between PE and mental health could possible explain this result. The negative association may also represent a pre-selection bias such that individuals with a high pre-existing psychopathological load may select specific sports. Other datasets may be more suitable to validate our findings and other approaches like Mendelian Randomization or randomized intervention designs can help to determine causal relationships in the future. Second, PE by definition (*50*) is a subset of PA that is planned, structured, and repetitive and has a final or an intermediate objective the improvement or maintenance of physical fitness. An important next step is to study the dose-response relationships for other more casual and unorganized forms of PA like walking, gardening and cleaning. Since current PE is assessed by parental report, wearable sensor data (e.g., Fitbit) could help us more accurately assess youths’ PA. Besides, the current study did not include other dosage measures like intensity (e.g., heart rate), which also need to be incorporated into our future work. Third, our analyses were basically cross-sectional. Longitudinal studies are needed to validate if current findings are consistent in the later stages of adolescence and explore how mental health changes with PE and brain development during this key transition period from adolescence to young adulthood. Finally, our genetic analyses only included youths of European ancestry as with numerous population genetics studies, which may restrict the generalization to non-European population. Multi-ancestry studies in the future are necessary to contribute to narrow health disparity in underrepresented populations (*51*).

In summary, the current results have important implications for both, public and individual mental health interventions. In particular the mitigating effects of exercise in adolescents with an increased genetic risk for mental health problems underscore that encouragement and opportunities to engage in exercise can promote resilience and in turn attenuate the vulnerability for mental disorders in later life. Taken together, these findings promote the application of personalized exercise-based interventions to improve mental health in adolescence and can inform individualized interventions that account for the interest of the adolescence, age and individual risk profile.

## Materials and Methods

### Participants

Participants are adolescents aged 9-10 years at baseline (n=11,878) from the USA recruited for the ABCD study (*25*) baseline assessment (Release 4.0), which is a multi-site, longitudinal and population-representative cohort. This cohort collects detailed data on physical and mental health, cognition, lifestyle, as well as genetic and neuroimaging data at baseline and follow-up. Demographic information of the full ABCD sample are provided in Table 1.

**Table 1.**
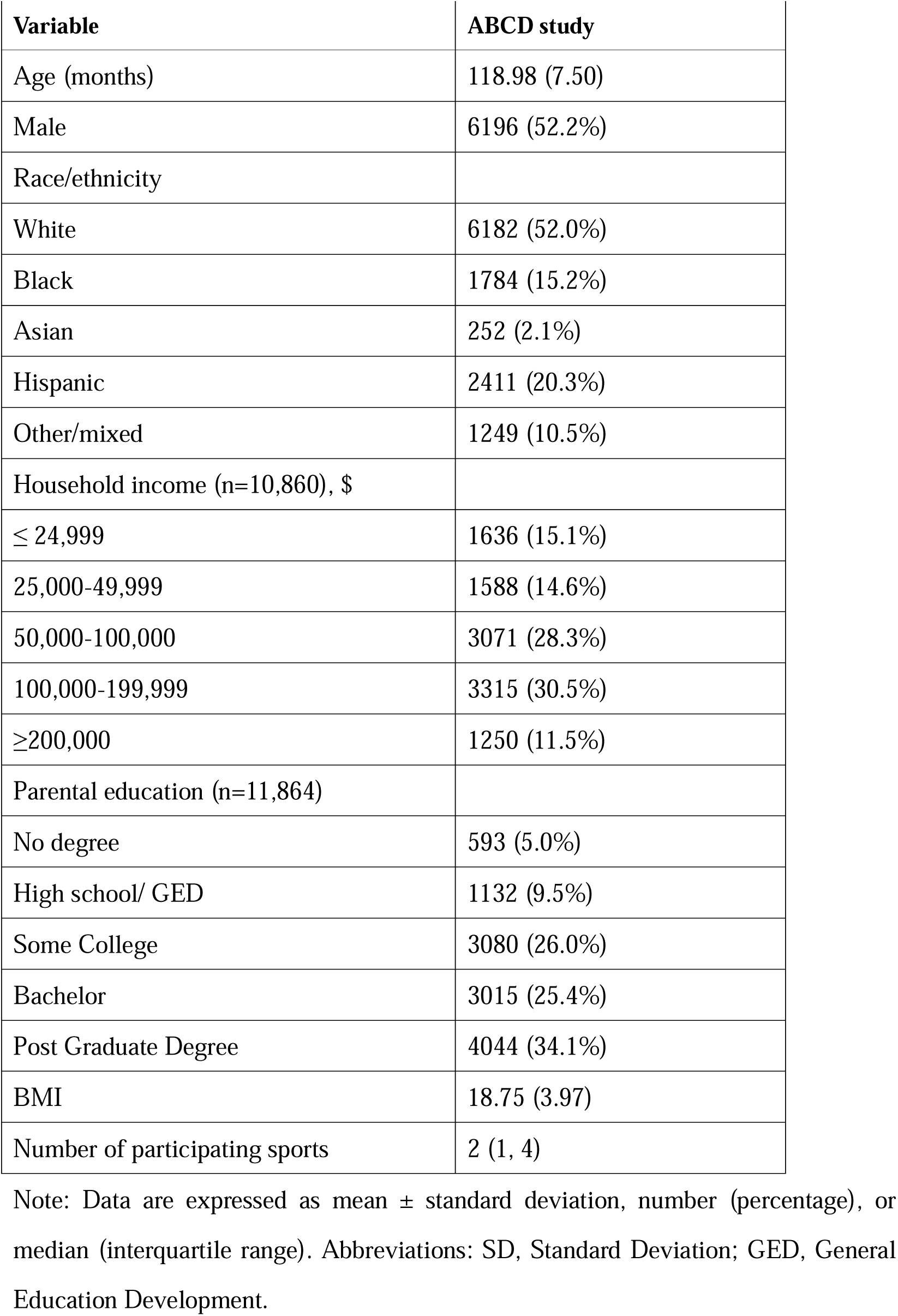
Demographics of the ABCD full sample (n=11 878).

**Table 2.** Demographics (sex and race) of 15 physical exercises.

|  | **Number** | **Sex (%)** |  | **Race (%)** |  |  |  |
| --- | --- | --- | --- | --- | --- | --- | --- |
|  | **Total**<br>**(n=11876)** | **Male**<br>**(n=6196)** | **Female**<br>**(n=5682)** | **White**<br>**(n=6182)** | **Black**<br>**(n=1784)** | **Hispanic**<br>**(n=2411)** | **Asian**<br>**(n=252)** |
| **Any** | 10032 (84.5) | 5310 (85.7) | 4722 (83.1) | 5612 (90.8) | 1243 (69.7) | 1917 (79.5) | 224 (88.9) |
| **ballet, dance** | 2994 (25.2) | 294 (4.7) | 2700 (47.5) | 1670 (27) | 381 (21.4) | 553 (22.9) | 66 (26.2) |
| **baseball,**<br>**softball** | 3220 (27.1) | 2305 (37.2) | 915 (16.1) | 2173 (35.2) | 245 (13.7) | 473 (19.6) | 34 (13.5) |
| **basketball** | 3136 (26.4) | 2202 (35.5) | 934 (16.4) | 1739 (28.1) | 541 (30.3) | 442 (18.3) | 64 (25.4) |
| **climbing** | 413 (3.5) | 225 (3.6) | 188 (3.3) | 254 (4.1) | 61 (3.4) | 48 (2) | 10 (4) |
| **football** | 1384 (11.7) | 1302 (21) | 82 (1.4) | 642 (10.4) | 394 (22.1) | 192 (8) | 10 (4) |
| **gymnastics** | 2635 (22.2) | 516 (8.3) | 2119 (37.3) | 1719 (27.8) | 199 (11.2) | 376 (15.6) | 66 (26.2) |
| **horseback**<br>**riding, polo** | 458 (3.9) | 125 (2) | 333 (5.9) | 317 (5.1) | 29 (1.6) | 66 (2.7) | 9 (3.6) |
| **ice/inline**<br>**skating** | 687 (5.8) | 245 (4) | 442 (7.8) | 437 (7.1) | 42 (2.4) | 117 (4.9) | 30 (11.9) |
| **martial arts** | 2076 (17.5) | 1410 (22.8) | 666 (11.7) | 1181 (19.1) | 176 (9.9) | 389 (16.1) | 65 (25.8) |
| **skateboarding** | 497 (4.2) | 366 (5.9) | 131 (2.3) | 250 (4) | 51 (2.9) | 125 (5.2) | 10 (4) |
| **skiing,**<br>**snowboarding** | 961 (8.1) | 535 (8.6) | 426 (7.5) | 798 (12.9) | 13 (0.7) | 48 (2) | 20 (7.9) |
| **soccer** | 4868 (41) | 2960 (47.8) | 1908 (33.6) | 3111 (50.3) | 295 (16.5) | 873 (36.2) | 84 (33.3) |
| **swimming,**<br>**water polo** | 3846 (32.4) | 1905 (30.7) | 1941 (34.2) | 2248 (36.4) | 414 (23.2) | 664 (27.5) | 126 (50) |

Note: The percentage in the bracket is the percentage of the number of each subgroup in the total number of each column.

### Measurements of physical exercise

For each kind of PE, the parents provided information on whether their child has ever participated in this PE continuously for 4 months or more. For those whose answer is ‘yes’, they were further asked about (1) for how many years, (2) how many months per year, (3) how many days per week and (4) how many minutes per session their child participated in this PE. Further, to estimate the total amount of each PE undertaken each week, we multiplied duration per session by days per week as the total amount (hours per week) for each PE type. We also approximately calculated the age of initiating each kind of PE by subtracting the cumulative years of participation from the age at baseline.

### Assessment of mental health

The Child Behavior Checklist (CBCL) (*52*) is an extensively validated and utilized parent-reported questionnaire to measure behavioral and emotional problems in school-aged children. CBCL in ABCD includes two empirically-derived broadband scales representing Internalizing and Externalizing Problems, one Total problems score, some syndrome scales like Anxious/Depressed, Thought Problems, Aggressive Behavior and some DSM-oriented scales like Attention Deficit/Hyperactivity Problems and Obsessive-Compulsive Problems. To align with the dimensional models of psychopathology (*53, 54*), which usually include one general psychopathology factor (‘p’ factor) as well as externalizing, internalizing and thought disorders, we used CBCL Externalizing Problems, Internalizing Problems, Thought Problems and Total Problems as primary outcomes.

### Structural and resting-state functional MRI

Participants completed a high-resolution T1-weighted structural MRI scan (1-mm isotropic voxels, TI=1060ms) and resting-state functional MRI scan (2.4 mm isotropic, TR=800ms, TE=30ms) on 3T scanners (Siemens Prisma, Phillips and GE 750). The full details of the image acquisition protocol were previously described in Casey et al. (*55*) and the detailed procedures of preprocessing and post-processing for structural MRI (sMRI) and resting-state functional MRI (rs-fMRI) data have been described in Hagler et al. (*56*). Morphometric measures include cortical thickness (CT), area (CA), volume (CV) and subcortical volume (subCV). Cortical morphometric measures were mapped to 34 cortical parcellations per hemisphere based on Desikan-Killiany Atlas (*57*) and subcortical measures were derived from FreeSurfer automatic segmentation (*58*). Within- and between-network resting-state functional connectivity for 12 predefined resting state networks was extracted by averaging all connections between ROIs assigned to given networks of the Gordon atlas (*59*). Average correlations between each network and each subcortical gray matter ROI (region of interest) were also calculated. We followed the recommended inclusion criterion for sMRI and rs-fMRI quality control in the ABCD 4.0 release and included 11231 participants for sMRI data and 9373 participants for rs-MRI data.

### Genotyping and imputation

Saliva samples collected during the baseline visit were genotyped using the Smokescreen array (*60*). Please refer to the reference (*61*) for details of biospecimen collection. Before imputation, we applied standard quality control on genotype data: (1) remove genotype data from plate 461, which is recommended by ABCD official release note due to poor data quality (2) remove individuals with missing genetics (3) remove single nucleotide polymorphism (SNP) with Minor Allele Frequency (MAF) <5% or sample missing >20%, leaving 371,316 SNPs for imputation. Next, we selected 5807 European ancestry individuals based on genetic ancestry proportion factors calculated by ABCD (genetic_af_european > 0.95). Further we only retained 4706 genetically unrelated individuals with kinship coefficient<0.125, calculated by plink 2.0 (*62*). Imputation was performed on 4706 genetically unrelated European ancestry individuals using the Michigan Imputation Server (*63*) with hrc.r1.1.2016 reference panel and Eagle v2.4 phasing. Best guess conversion at a threshold of 0.9 was used to convert dosage files to plink binary PED files. After imputation, we excluded individuals with >10% missing rate and SNPs with low imputation quality, >5% missing rate, MAF <1%, or out of Hardy-Weinberg equilibrium violation (p>10^-6^), yielding 4,326,912 SNPs. In order to correct for population stratification, genetic principal component analysis was performed on these unrelated European individuals to derive first 10 genetic principal components (PCs).

### Polygenic risk scores

The polygenic risk score (PRS) characterizes an individual’s genetic predisposition to a disease or trait by aggregating the effect size of genetic variants in Genome-Wide Association Study (GWAS). To comprehensively quantify adolescents’ genetic risks of psychopathology, we selected 10 mental disorders or psychological traits with publicly available GWAS summary statistics, including schizophrenia (SCZ) (*26*), psychotic experiences (PSE) (*27*), Post-traumatic stress disorder (PTSD) (*28*), bipolar disorder (BIP) (*29*), major depressive disorder (MDD) (*30*), depression (*31*), neuroticism (*31*), Obsessive-compulsive disorder (OCD) (*32*), Attention-Deficit / Hyperactivity Disorder (ADHD) (*33*) and risk tolerance (*34*). We used PRS-CS (*64*) and 1000 Genomes Project Phase 3 European population as linkage disequilibrium (LD) reference panel to calculate 9 PRSs for above mental disorders or psychological traits in 4706 unrelated European ancestry adolescents. Standardized PRSs were used in following analyses.

### Statistical analyses

We first used linear mixed models (LMM) implemented in R *lme4* (*65*) package to examine the relationships between previous participation of each kind of PE and four CBCL scales for dimensional psychopathology. We used a two-side p<0.001 significance threshold. For those PEs with significant beneficial effects on psychopathology, we further examined the relationships between six other dosage measures of PEs and dimensional psychology. Secondary analyses also examined the associations with other 14 CBCL scales. LMM included covariates for age (except for age of starting exercising), sex, race, household income, parental education, pubertal level, body max index (BMI), and random effects for family nested within site. False discovery rate (FDR) correction was used in secondary analyses to correct for 14 comparisons. Further, to investigate the possibly nonlinear relationships, we also compared the differences in mental health burden between lower level group and other groups and further examined if there existed linear trend relationships across groups. Months per year is categorized as 1-3 months, 4-6 months, 7-9 months and 10-12 months. Days per week is categorized as ≤1 day, 2-3 days, 4-5 days and >5 days. The duration per session of each PE is divided into<30 min, 30-60 min, 60-90 min, 90-120 min and ≥120 min. The amount per week of each PE is divided into <1 hour, 1-2 hours, 2-3 hours, 3-4 hours and >4 hours. Age of starting exercising is divided into toddler (<3 years), preschooler (3-5 years), middle childhood (6-8 years) and early adolescence (9-10 years). Lowest dose level or latest age was set as reference group To investigate the neural correlates of PE with beneficial effects, we examined the associations between 7 dosage measures of PEs and brain morphology (CT, CA, CV and subCV) and functional network measures (resting-state functional connectivity between and within cortical networks, and between cortical networks and subcortical ROIs) using LMM. LMM additionally adjusted MRI scanners and intracranial volume for sMRI or mean framewise displacement for rs-fMRI. FDR correction was used to correct for multiple comparison. Next, we explored whether PE-related neural correlates mediated the beneficial effects of PE on mental health. The mediation analyses were performed using the Mediation toolbox (*66*) in MATLAB with 10,000 bootstraps after regressing out same covariates as above. Finally, to explore how the interactions between genetic risks and PEs influence mental health, we also examined the effects of interactions between PRS and exercise dosage on mental health. Age, sex, household income, parental education, BMI, pubertal level, top 10 PCs and study sites were adjusted. FDR correction was used to correct for multiple comparisons.

## Data Availability

The data from ABCD study is available by request (See https://abcdstudy.org/scientists/data-sharing/).

## Acknowledgements

JZ was supported by Science and Technology Innovation 2030 - Major Projects (Grant No.2021ZD0200204), Science and Technology Innovation 2030 - Brain Science and Brain-Inspired Intelligence Project (Grant No. 2021ZD0200204), Shanghai Municipal Science and Technology Major Project (No.2018SHZDZX01), ZJLab, and NSFC 61973086. JF was supported by the 111 Project (No. B18015), the key project of Shanghai Science and Technology (No. 16JC1420402), National Key R&D Program of China (No. 2018YFC1312900), and National Natural Science Foundation of China (NSFC 91630314).

## Notes

### Competing Interest Statement

The authors have declared no competing interest.

## References

1. M. Hirvensalo, T. Lintunen, Life-course perspective for physical activity and sports participation. European Review of Aging and Physical Activity 8, 13–22 (2011).

2. K. Corder et al., Change in physical activity from adolescence to early adulthood: a systematic review and meta-analysis of longitudinal cohort studies. British journal of sports medicine 53, 496–503 (2019).

3. R. Telama, Tracking of physical activity from childhood to adulthood: a review. Obesity facts 2, 187–195 (2009).

4. A. Caspi et al., Longitudinal Assessment of Mental Health Disorders and Comorbidities Across 4 Decades Among Participants in the Dunedin Birth Cohort Study. Jama Netw Open 3, e203221–e203221 (2020).

5. R. C. Kessler et al., Lifetime prevalence and age-of-onset distributions of DSM-IV disorders in the National Comorbidity Survey Replication. Arch Gen Psychiatry 62, 593–602 (2005).

6. C. P. McDowell, R. K. Dishman, B. R. Gordon, M. P. Herring, Physical activity and anxiety: a systematic review and meta-analysis of prospective cohort studies. American journal of preventive medicine 57, 545–556 (2019).

7. A. Kandola et al., Moving to beat anxiety: epidemiology and therapeutic issues with physical activity for anxiety. Current psychiatry reports 20, 1–9 (2018).

8. F. B. Schuch et al., Physical activity and incident depression: a meta-analysis of prospective cohort studies. American Journal of Psychiatry 175, 631–648 (2018).

9. S. Kvam, C. L. Kleppe, I. H. Nordhus, A. Hovland, Exercise as a treatment for depression: a meta-analysis. Journal of affective disorders 202, 67–86 (2016).

10. B. R. Belcher et al., The roles of physical activity, exercise, and fitness in promoting resilience during adolescence: effects on mental well-being and brain development. Biological Psychiatry: Cognitive Neuroscience and Neuroimaging 6, 225–237 (2021).

11. G. Ashdown-Franks et al., Exercise as medicine for mental and substance use disorders: a meta-review of the benefits for neuropsychiatric and cognitive outcomes. Sports Medicine 50, 151–170 (2020).

12. J. Kou et al., Anxiolytic effects of chronic intranasal oxytocin on neural responses to threat are dose-frequency dependent. Psychotherapy and Psychosomatics 91, 253–264 (2022).

13. J. Kou et al., A randomized trial shows dose-frequency and genotype may determine the therapeutic efficacy of intranasal oxytocin. Psychological Medicine 52, 1959–1968 (2022).

14. S. R. Chekroud et al., Association between physical exercise and mental health in 1· 2 million individuals in the USA between 2011 and 2015: a cross-sectional study. The Lancet Psychiatry 5, 739–746 (2018).

15. K. Kunitoki et al., Youth Team Sports Participation Associates With Reduced Dimensional Psychopathology Through Interaction With Biological Risk Factors. Biological Psychiatry Global Open Science, (2023).

16. L. S. Gorham, T. Jernigan, J. Hudziak, D. M. Barch, Involvement in sports, hippocampal volume, and depressive symptoms in children. Biological Psychiatry: Cognitive Neuroscience and Neuroimaging 4, 484–492 (2019).

17. C. W. Cotman, N. C. Berchtold, L.-A. Christie, Exercise builds brain health: key roles of growth factor cascades and inflammation. Trends in neurosciences 30, 464–472 (2007).

18. M. W. Voss, C. Vivar, A. F. Kramer, H. van Praag, Bridging animal and human models of exercise-induced brain plasticity. Trends in cognitive sciences 17, 525–544 (2013).

19. J. Firth et al., Effect of aerobic exercise on hippocampal volume in humans: a systematic review and meta-analysis. Neuroimage 166, 230–238 (2018).

20. K. I. Erickson, R. L. Leckie, A. M. Weinstein, Physical activity, fitness, and gray matter volume. Neurobiology of aging 35, S20–S28 (2014).

21. Q. Yu et al., Cognitive benefits of exercise interventions: an fMRI activation likelihood estimation meta-analysis. Brain Structure and Function 226, 601–619 (2021).

22. M. Rodriguez-Ayllon et al., Role of physical activity and sedentary behavior in the mental health of preschoolers, children and adolescents: a systematic review and meta-analysis. Sports medicine 49, 1383–1410 (2019).

23. C. M. Stillman, I. Esteban-Cornejo, B. Brown, C. M. Bender, K. I. Erickson, Effects of exercise on brain and cognition across age groups and health states. Trends in neurosciences 43, 533–543 (2020).

24. D. M. Barch et al., Demographic, physical and mental health assessments in the adolescent brain and cognitive development study: Rationale and description. Developmental cognitive neuroscience 32, 55–66 (2018).

25. D. M. Barch et al., Demographic, physical and mental health assessments in the adolescent brain and cognitive development study: Rationale and description. Dev Cogn Neurosci 32, 55–66 (2018).

26. V. Trubetskoy et al., Mapping genomic loci implicates genes and synaptic biology in schizophrenia. Nature 604, 502–508 (2022).

27. S. E. Legge et al., Association of genetic liability to psychotic experiences with neuropsychotic disorders and traits. JAMA psychiatry 76, 1256–1265 (2019).

28. C. M. Nievergelt et al., International meta-analysis of PTSD genome-wide association studies identifies sex-and ancestry-specific genetic risk loci. Nature communications 10, 1–16 (2019).

29. N. Mullins et al., Genome-wide association study of more than 40,000 bipolar disorder cases provides new insights into the underlying biology. Nature genetics 53, 817–829 (2021).

30. N. R. Wray et al., Genome-wide association analyses identify 44 risk variants and refine the genetic architecture of major depression. Nature genetics 50, 668–681 (2018).

31. M. Nagel et al., Meta-analysis of genome-wide association studies for neuroticism in 449,484 individuals identifies novel genetic loci and pathways. Nature genetics 50, 920–927 (2018).

32. P. D. Arnold et al., Revealing the complex genetic architecture of obsessive-compulsive disorder using meta-analysis. Molecular psychiatry 23, 1181–1181 (2018).

33. D. Demontis et al., Discovery of the first genome-wide significant risk loci for attention deficit/hyperactivity disorder. Nature genetics 51, 63–75 (2019).

34. R. Karlsson Linnér et al., Genome-wide association analyses of risk tolerance and risky behaviors in over 1 million individuals identify hundreds of loci and shared genetic influences. Nature genetics 51, 245–257 (2019).

35. M. D. Hoffmann, J. D. Barnes, M. S. Tremblay, M. D. Guerrero, Associations between organized sport participation and mental health difficulties: Data from over 11,000 US children and adolescents. PLoS one 17, e0268583 (2022).

36. A. N. Kaczkurkin et al., Approaches to defining common and dissociable neurobiological deficits associated with psychopathology in youth. Biological psychiatry 88, 51–62 (2020).

37. G. Cooney, Exercise and mental health: a complex and challenging relationship. The Lancet Psychiatry 5, 692–693 (2018).

38. S. B. Harvey et al., Exercise and the prevention of depression: results of the HUNT cohort study. American Journal of Psychiatry 175, 28–36 (2018).

39. M. Rodriguez-Ayllon et al., Neurobiological, psychosocial, and behavioral mechanisms mediating associations between physical activity and psychiatric symptoms in youth in the Netherlands. JAMA psychiatry 80, 451–458 (2023).

40. N. M. Collins, F. Cromartie, S. Butler, J. Bae, Effects of early sport participation on self-esteem and happiness. The sport journal 20, 1–20 (2018).

41. L. T. Muftuler et al., Cortical and subcortical changes in typically developing preadolescent children. Brain research 1399, 15–24 (2011).

42. M. Rodriguez-Ayllon et al., Associations of physical activity and screen time with white matter microstructure in children from the general population. Neuroimage 205, 116258 (2020).

43. D. H. Weissman, K. Roberts, K. Visscher, M. Woldorff, The neural bases of momentary lapses in attention. Nature neuroscience 9, 971–978 (2006).

44. S. Vossel, J. J. Geng, G. R. Fink, Dorsal and ventral attention systems: distinct neural circuits but collaborative roles. The Neuroscientist 20, 150–159 (2014).

45. R. H. Kaiser, J. R. Andrews-Hanna, T. D. Wager, D. A. Pizzagalli, Large-scale network dysfunction in major depressive disorder: a meta-analysis of resting-state functional connectivity. JAMA psychiatry 72, 603–611 (2015).

46. C. M. Sylvester et al., Functional network dysfunction in anxiety and anxiety disorders. Trends in neurosciences 35, 527–535 (2012).

47. B. L. Hankin, B. E. Gibb, J. R. Abela, K. Flory, Selective attention to affective stimuli and clinical depression among youths: role of anxiety and specificity of emotion. Journal of abnormal psychology 119, 491 (2010).

48. P. S. Tandon et al., Socioeconomic inequities in youth participation in physical activity and sports. International Journal of Environmental Research and Public Health 18, 6946 (2021).

49. S. M. P. Pereira, M.-C. Geoffroy, C. Power, Depressive symptoms and physical activity during 3 decades in adult life: bidirectional associations in a prospective cohort study. JAMA psychiatry 71, 1373–1380 (2014).

50. C. J. Caspersen, K. E. Powell, G. M. Christenson, Physical activity, exercise, and physical fitness: definitions and distinctions for health-related research. Public health reports 100, 126 (1985).

51. A. R. Martin et al., Clinical use of current polygenic risk scores may exacerbate health disparities. Nature genetics 51, 584–591 (2019).

52. T. M. Achenbach, The Achenbach system of empirically based assessment (ASEBA): Development, findings, theory, and applications. (University of Vermont, Research Center for Children, Youth, & Families, 2009).

53. A. Caspi et al., The p Factor: One General Psychopathology Factor in the Structure of Psychiatric Disorders? Clin Psychol Sci 2, 119–137 (2014).

54. A. Caspi, T. E. Moffitt, All for One and One for All: Mental Disorders in One Dimension. American Journal of Psychiatry 175, 831–844 (2018).

55. B. J. Casey et al., The Adolescent Brain Cognitive Development (ABCD) study: Imaging acquisition across 21 sites. Dev Cogn Neurosci 32, 43–54 (2018).

56. D. J. Hagler Jr et al., Image processing and analysis methods for the Adolescent Brain Cognitive Development Study. Neuroimage 202, 116091 (2019).

57. R. S. Desikan et al., An automated labeling system for subdividing the human cerebral cortex on MRI scans into gyral based regions of interest. Neuroimage 31, 968–980 (2006).

58. B. Fischl et al., Whole brain segmentation: automated labeling of neuroanatomical structures in the human brain. Neuron 33, 341–355 (2002).

59. E. M. Gordon et al., Generation and evaluation of a cortical area parcellation from resting-state correlations. Cerebral cortex 26, 288–303 (2016).

60. J. W. Baurley, C. K. Edlund, C. I. Pardamean, D. V. Conti, A. W. Bergen, Smokescreen: a targeted genotyping array for addiction research. BMC Genomics 17, 145 (2016).

61. K. A. Uban et al., Biospecimens and the ABCD study: Rationale, methods of collection, measurement and early data. Dev Cogn Neurosci 32, 97–106 (2018).

62. C. C. Chang et al., Second-generation PLINK: rising to the challenge of larger and richer datasets. Gigascience 4, 7 (2015).

63. S. Das et al., Next-generation genotype imputation service and methods. Nat Genet 48, 1284–1287 (2016).

64. T. Ge, C.-Y. Chen, Y. Ni, Y.-C. A. Feng, J. W. Smoller, Polygenic prediction via Bayesian regression and continuous shrinkage priors. Nature communications 10, 1–10 (2019).

65. D. Bates, M. Mächler, B. Bolker, S. Walker, Fitting linear mixed-effects models using lme4. arXiv preprint arXiv:1406.5823, (2014).

66. T. D. Wager, M. L. Davidson, B. L. Hughes, M. A. Lindquist, K. N. Ochsner, Prefrontal-subcortical pathways mediating successful emotion regulation. Neuron 59, 1037–1050 (2008).

