## Supplemental Materials for "Dose-Response Relationships between Physical Exercises and Mental Health during Early Adolescence: an Investigation of the Underlying Neural and Genetic Mechanisms from the ABCD Study"

#### Recoding the dosage measures of physical exercise

Before regression, we recoded days per week and duration per session in ABCD into a more comparable numbers. For days per week, we recoded 8 (every 2 weeks) as 0.5, 9 (One day every month) as 0.25 and 10 (Less than one day per month) as 0.1. For duration per session, we recoded 5 (90 min) as 6, 6 (120 min) as 8, 7 (150 min) as 10, 8 (3 hours) as 12 and 9 (greater than 3 hours) as 14.

### Supplemental figures

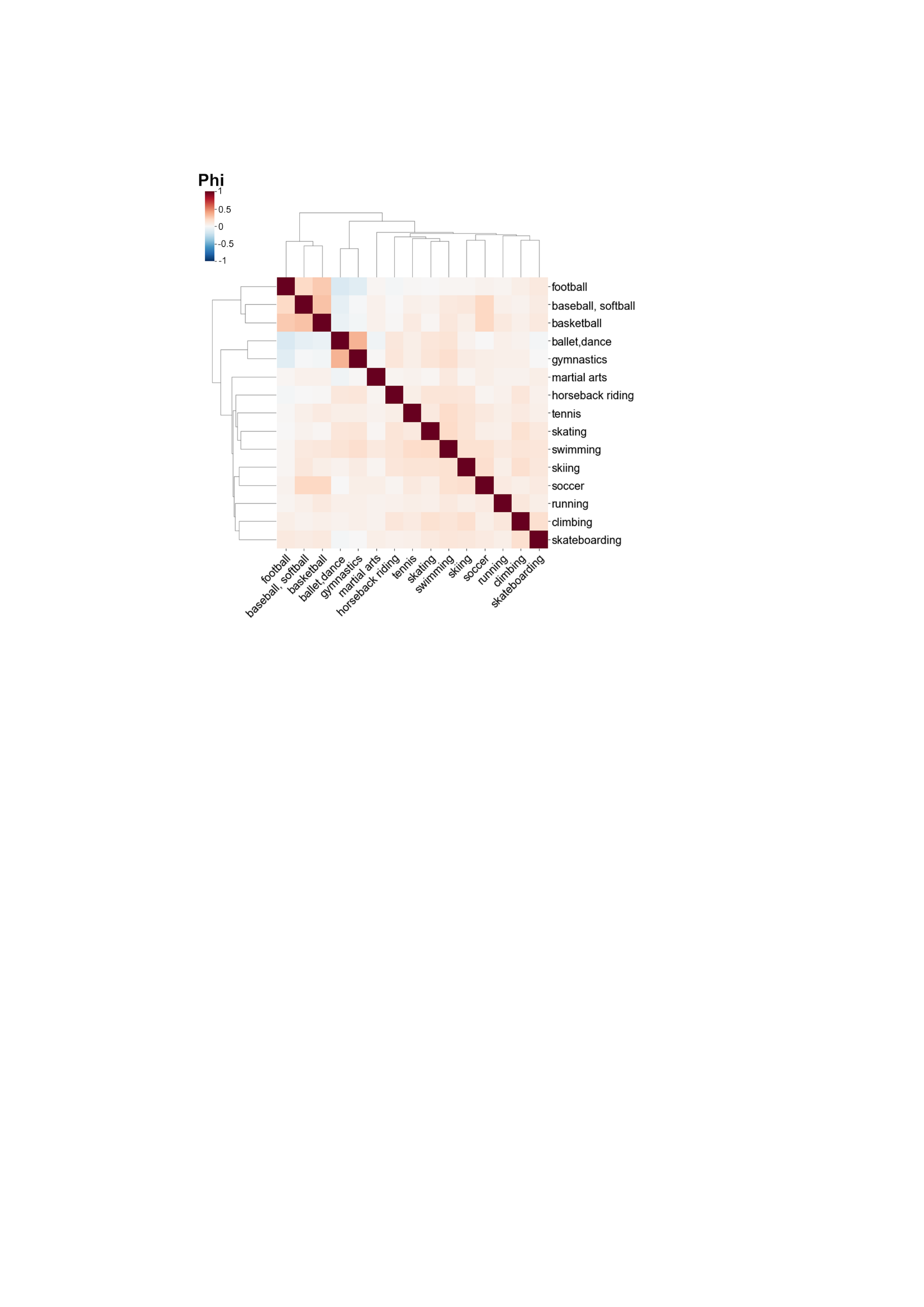

**Figure S1**. Correlations between participations of different physical exercises. The colorbar represent the Phi correlation coefficient. The dendrograms in X-axis and Y-axis showed the hierarchical clustering based on correlations between physical exercises.

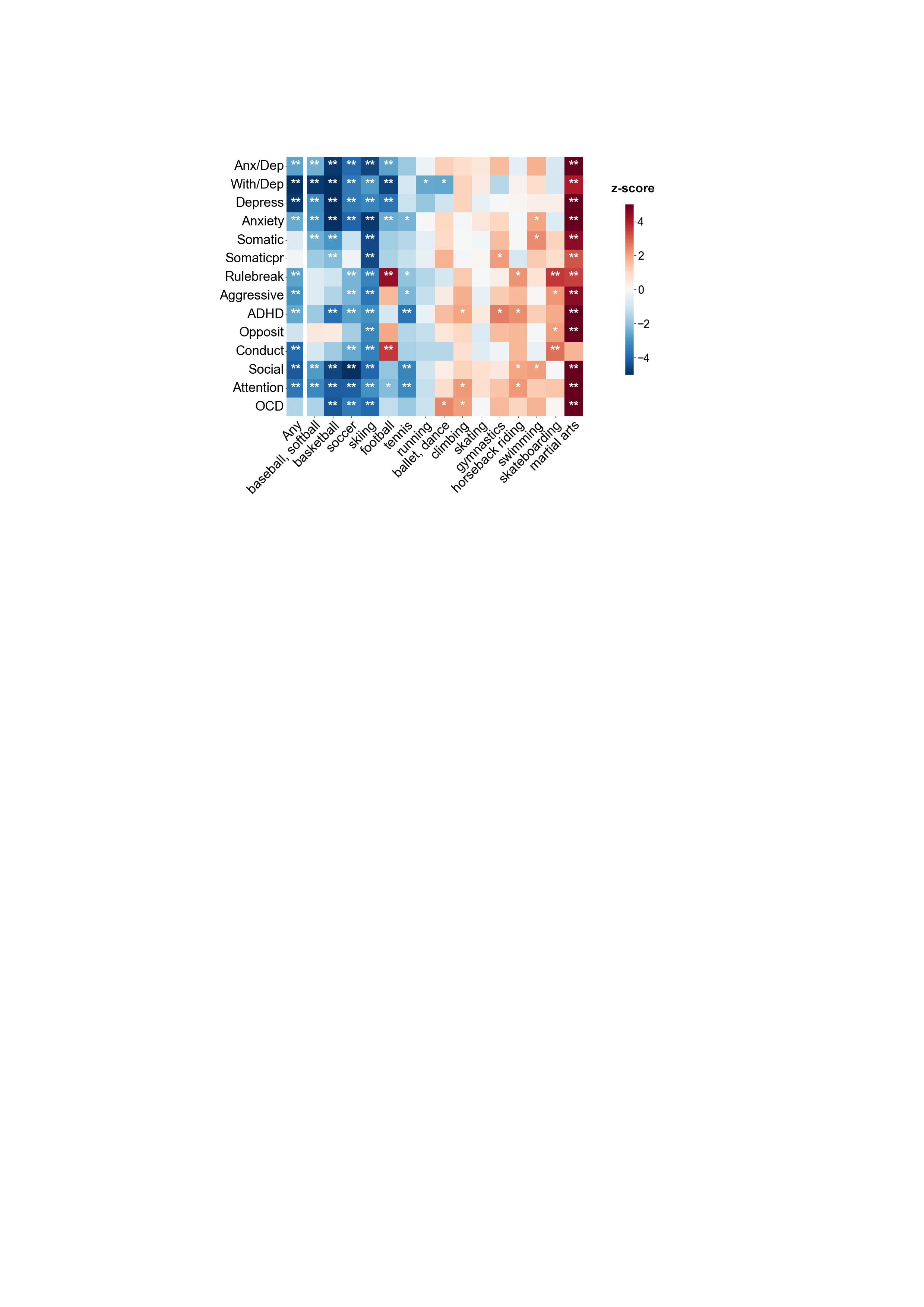

**Figure S2.** The relationships between previous participation of 15 kinds of PEs (as well as participation of any PEs) and CBCL subscales. The colorbar represents the z-score of the regression coefficient from LMM. *: p<0.05; **: P_FDR_<0.05.

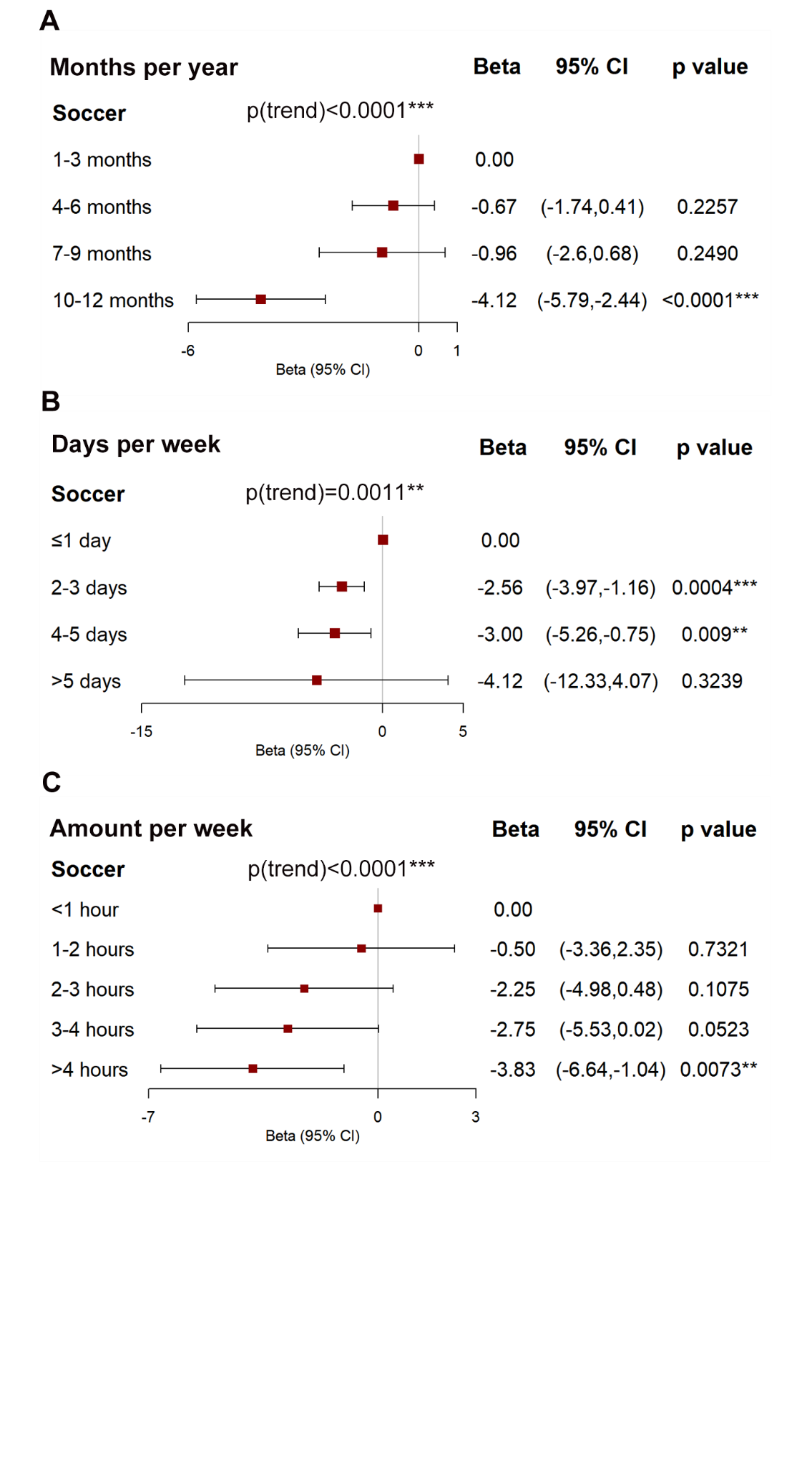

**Figure S3.** The associations between different levels of **(A)** months per year, (B) days per week and **(C)** amount per week of playing soccer and CBCL Total Problems. Betas were adjusted for sex, race, household income, parental education, pubertal level, body max index (BMI), and random effects for family nested within site. p(trend) means p value for linear trend test. The asterisks indicate: *, p<0.05; **, p<0.01; ***, p<0.001.

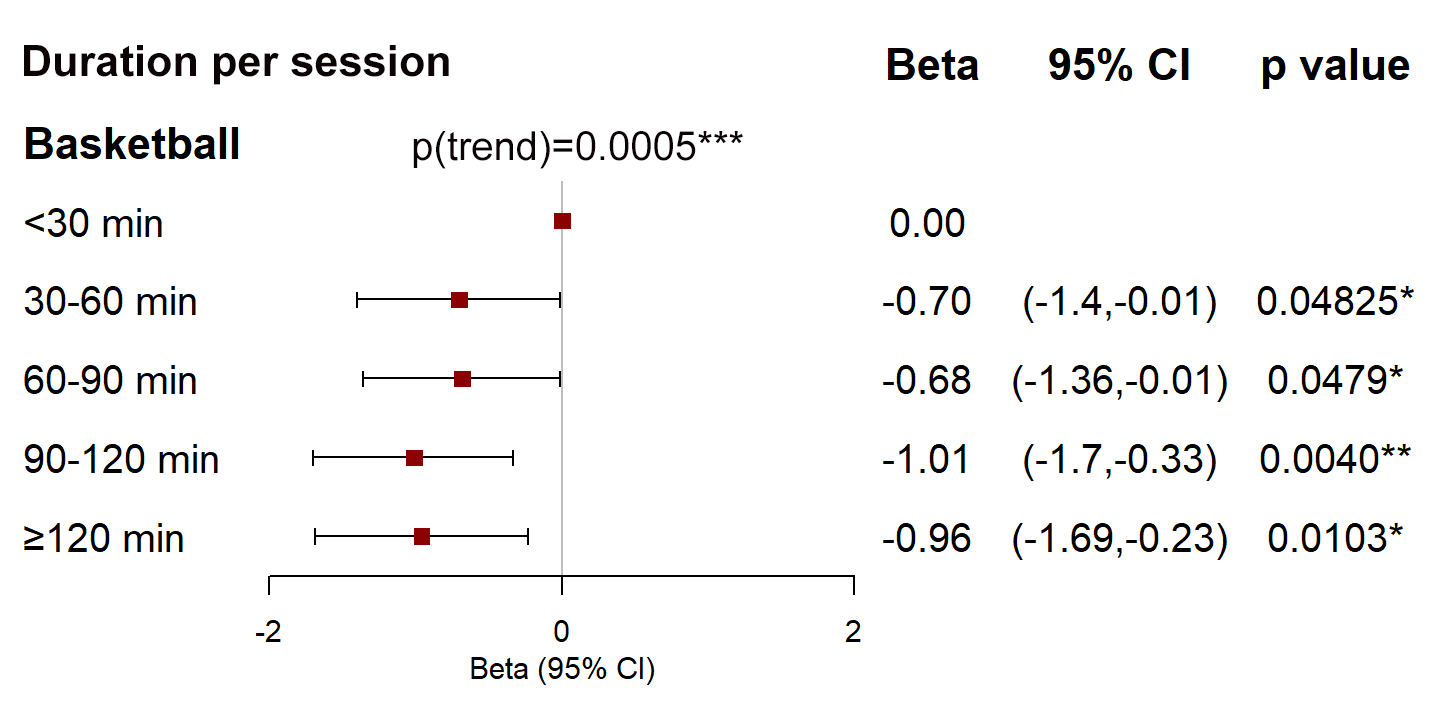

**Figure S4.** The associations between different groups of duration per session for playing basketball and CBCL Thought Problems. Betas were adjusted for sex, race, household income, parental education, pubertal level, body max index (BMI), and random effects for family nested within site. p(trend) means p value for linear trend test. *: p<0.05; **: p<0.01; ***: p<0.001.

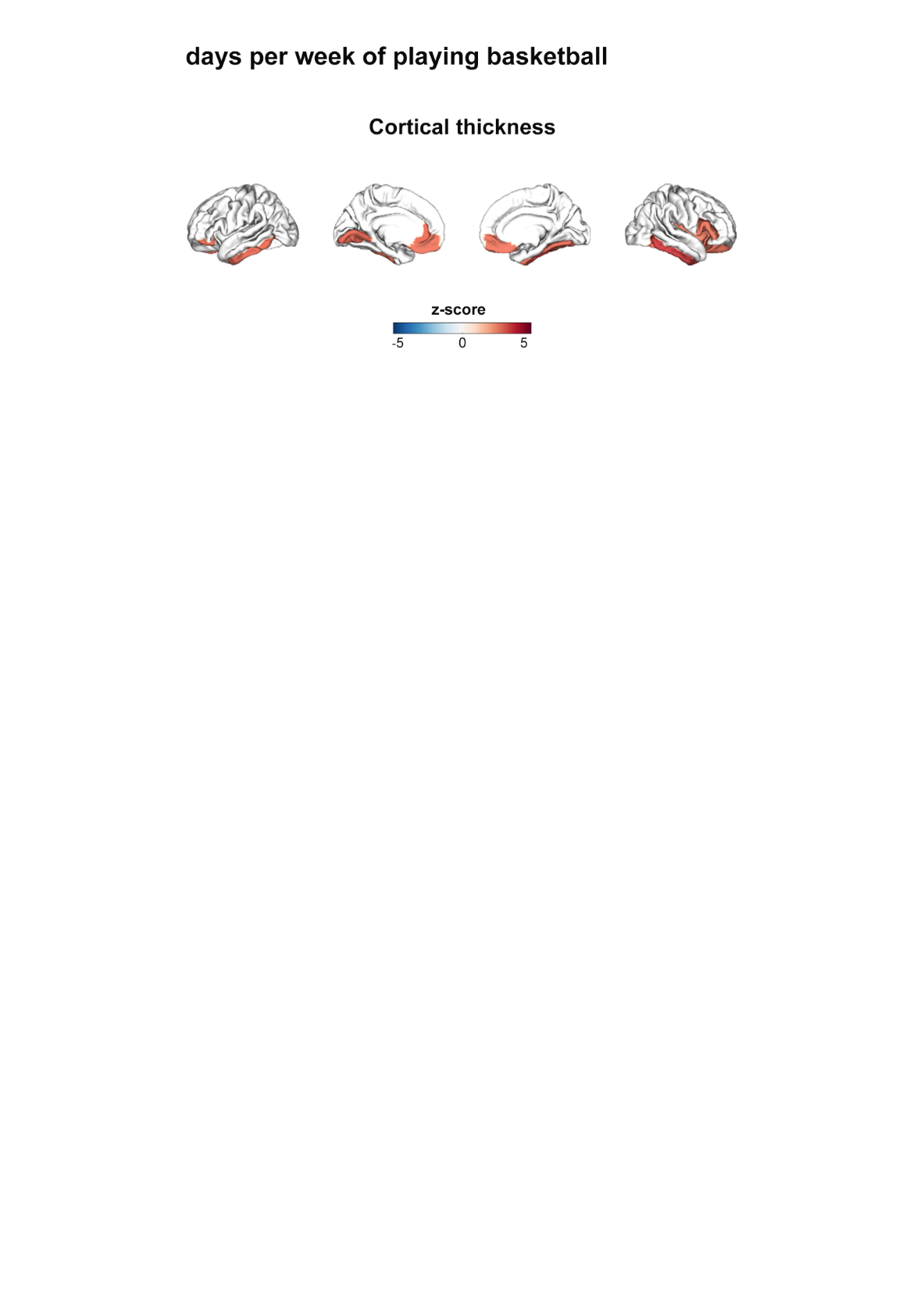

**Figure S5.** Cortical thickness significantly (P_FDR_<0.05) associated with days per week of playing basketball. The colorbar represent the z-score of the regression coefficient from LMM.

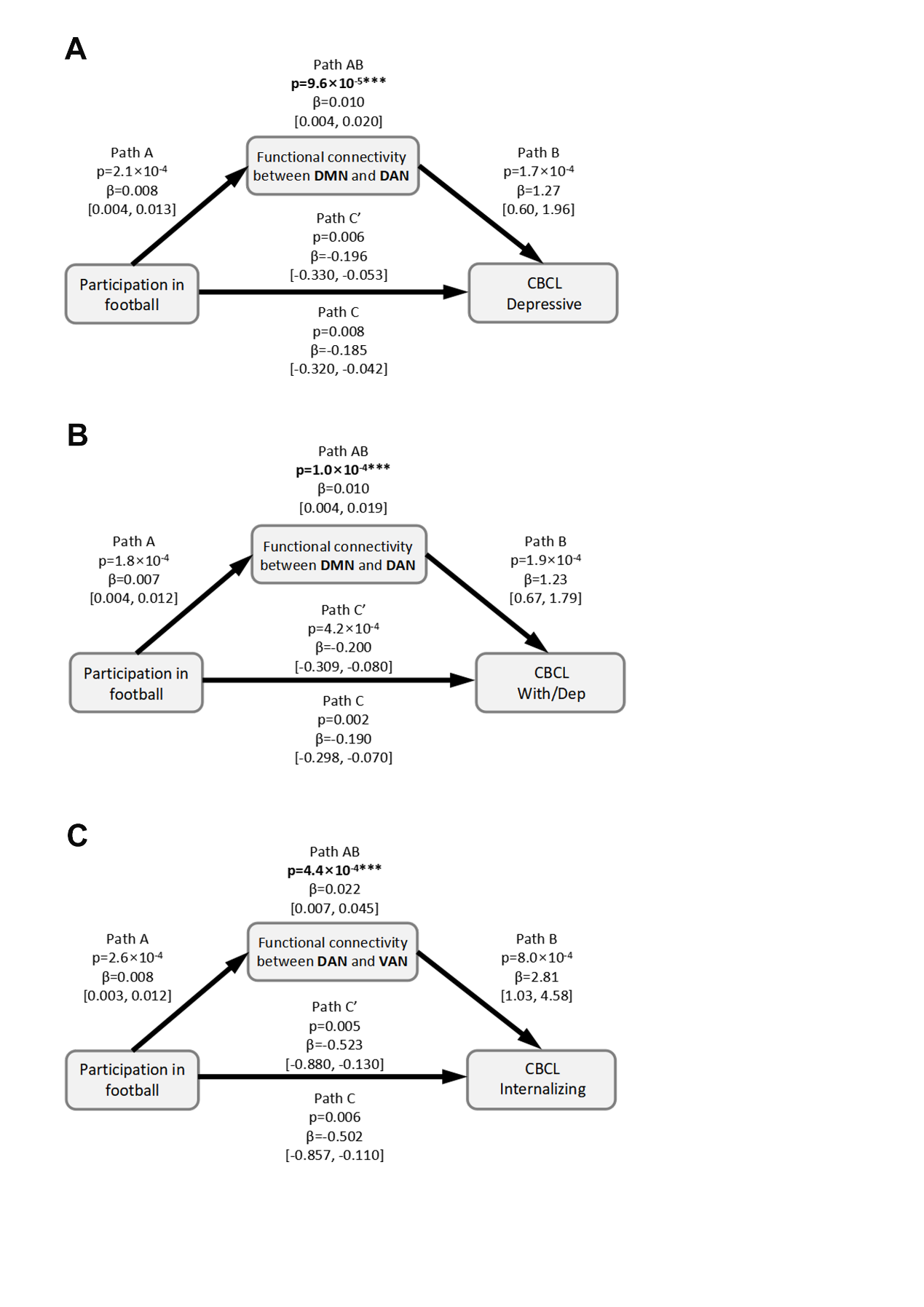

**Figure S6.** Mediation analysis. **(A)** Functional connectivity between default mode network (DMN) and dorsal attention network (DAN) mediates the beneficial effect of participation in football on CBCL Depressive Problems. **(B)** Functional connectivity between DMN and DAN mediates the effect of participation in football on CBCL Withdrawn/Depressed Problems. **(C)** Functional connectivity between DAN and VAN mediates the effect of participation in football on CBCL Internalizing Problems. The 95% confidence intervals are listed in the bracket. In **(A-C)**, Path AB represents the average causal mediation effects (ACME). Path C represents the total effect without mediator. Path C’ represents the direct effect accounting for the indirect effect of mediator. *: p<0.05; **: p<0.01; ***: p<0.001.

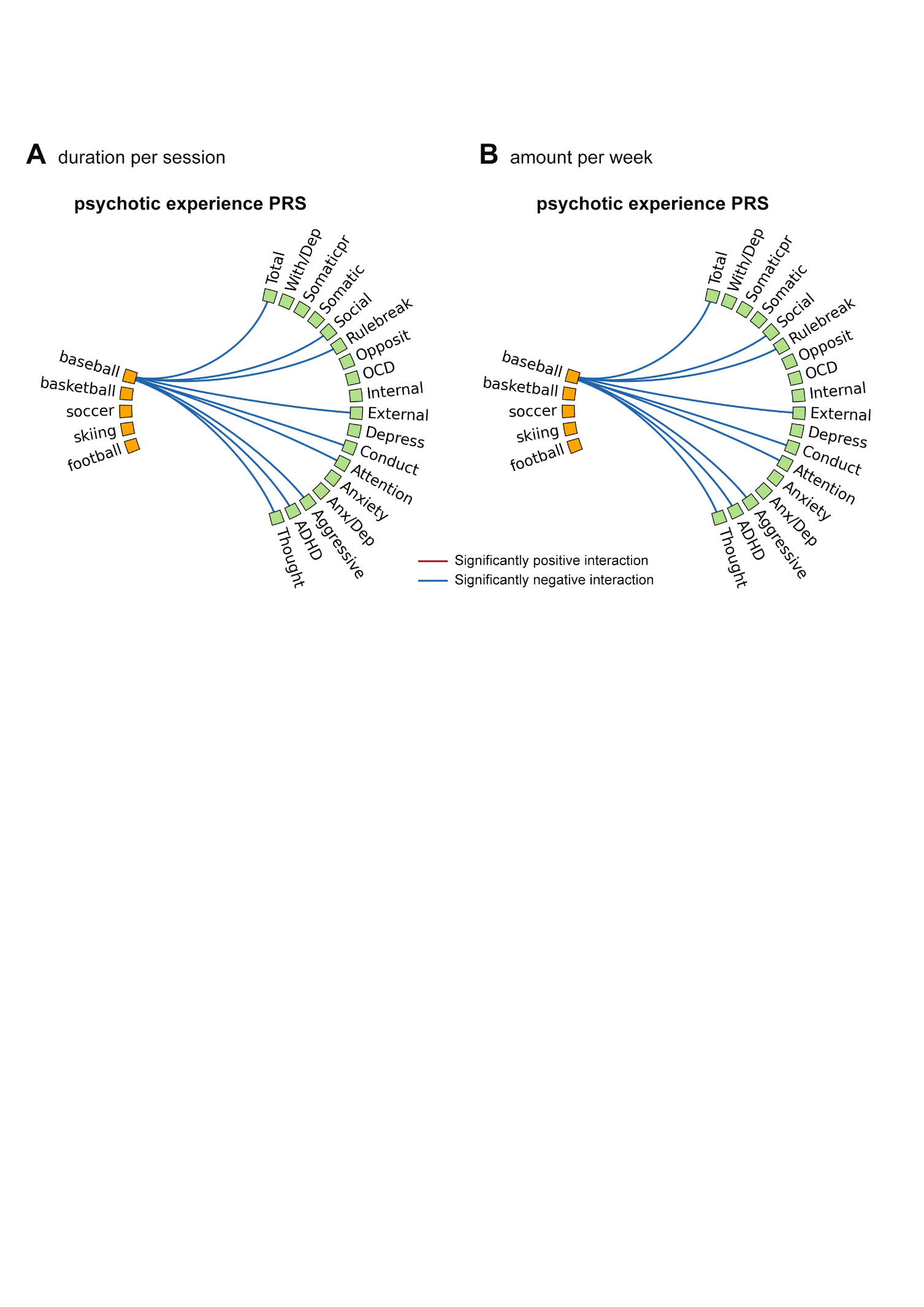

**Figure S7.** The effects of the interactions between PRS for psychotic experience and ‘beneficial’ physical exercises on mental health. The red line indicates significantly positive interaction between PRS for psychotic experience and physical exercises while the blue line indicates significantly negative interaction between PRS for psychotic experience and physical exercises.
